# Six-Minute Knee MRI: A Comparison of Novel Approaches for Accelerated Imaging

**DOI:** 10.1101/2025.02.11.25321839

**Authors:** Ananya Goyal, James MacKay, Matthew Petterson, Rianne van der Heijden, Kathryn Stevens, Min Yoon, Tie Liang, Akshay Chaudhari, Feliks Kogan

## Abstract

**Background:** Accelerated knee MRI protocols using deep learning (DL)-based reconstruction, 3D acquisitions, and parallel imaging can significantly reduce scan times. These methods enhance patient throughput and comfort while maintaining diagnostic quality. However, their clinical efficacy and diagnostic performance across various knee pathologies requires systematic evaluation.

**Purpose:** To evaluate the diagnostic performance and image quality of four accelerated knee MRI protocols (∼6 minutes) compared with the standard conventional 2D FSE protocol (∼15–30 minutes).

**Materials and Methods:** This prospective study enrolled adults with symptomatic knee pain between May 2021 and March 2022. All participants underwent five knee MRI protocols: a conventional 2D fast spin echo (FSE) protocol, a 2D FSE protocol with DL reconstruction, a 3D Cube protocol, a 3D quantitative double-echo steady-state (qDESS) protocol, and a thin-slice 2D FSE protocol. Five radiologists evaluated pathologies in joint tissues. Inter-reader and inter-method agreements were assessed using Gwet’s AC, with sensitivity and specificity calculated. Image diagnostic quality was evaluated using a Likert scale, and the Friedman was test used (p<0.05 indicative of statistically significant difference).

**Results:** A total of 32 participants were evaluated (14 men, aged 47±16 years). Of 160 total scans, 12 were excluded due to motion artifacts. Across pathologies, the 3D qDESS protocol had the highest inter-reader agreement for menisci (0.86) and cartilage (0.48), with sensitivities of 95% and 83%, respectively. The 2D DL protocol showed strong performance for bone marrow (sensitivity: 78%). Lastly, for effusion, both the 2D DL and 3D Cube protocols exhibited high inter-reader agreement (0.89 and 0.87, respectively). Diagnostic image quality scores exceeded were diagnostically acceptable in 95% of evaluations for menisci, bone marrow, and ligaments.

**Conclusion:** The 2D DL, 3D Cube, and 3D qDESS protocols matched the diagnostic performance of the conventional protocol while reducing scan times to six minutes, improving workflow and patient experience.

## 1. Introduction

Magnetic Resonance Imaging (MRI) is often the modality of choice for diagnostic musculoskeletal imaging, due to its excellent soft-tissue sensitivity with multiple contrasts and high resolution. It is commonly used to assess knee anatomy and pathology in a wide range of tissues, such as cartilage, bone, menisci, synovium, muscles, tendons, and ligaments. The number of knee MRI examinations continues to rise, with around 70,00 scans being conducted annually between 2019 and 2023 at a tertiary center like ours.

Due to the rising demand for knee MRI exams, it is imperative to reduce knee MRI scan times. Conventional clinical knee MRI protocols generally comprise of two-dimensional (2D) fast spin echo (FSE) sequences with high in-plane spatial resolution and multiple contrasts, such as T2-weighting, T1-weighting, proton-density (PD)-weighting, acquired in orthogonal scan planes^1^. However, conventional protocols have some limitations in the form of inefficient acquisitions, thick slices with slice spacing limiting visualization of subtle pathology, and potential motion artifacts due to long scan times. Despite using acceleration techniques and improved coils, such protocols are signal-to-noise ratio (SNR)-inefficient with total scan times of 15-30 mins.

Numerous approaches have been proposed to improve the value of knee imaging by reducing protocol times to less than six minutes and potentially adding diagnostic potential through multiplanar reformats (MPRs), new contrasts, or quantitative analysis. By utilizing improved acceleration techniques, such as parallel imaging acquisition and deep learning (DL)-based image denoising, both 2D and three-dimensional (3D) sequences can be shortened while preserving both image and diagnostic quality^2,3^. Furthermore, 3D sequences allow for MPRs, which may reduce the need for multiple image acquisitions in other planes^4,5^. However, a systematic comparison between these different protocols for accelerated knee MRI has not been conducted.

To address the aforementioned challenges, the purpose of this prospective, multi-reader study was to compare four accelerated knee MRI protocols against the reference conventional clinical protocol for assessment of internal derangements of the knee. The four protocols include combinations of 2D and 3D sequences, incorporating modern DL reconstruction, quantitative data, and increased resolution and SNR. Specifically, we aim to conduct a head-to-head comparison of the protocols’ diagnostic utility and quality with respect to fluid sensitivity and ligament, meniscus, and cartilage pathology of the four protocol types with that of the standard clinical knee MRI protocol.

## 2. Materials and Methods

### 2.1. Participants

This prospective, single-center study with MRI evaluation of five readers from three centers was approved by the Institutional Review Board. All participants gave written informed consent prior to study participation and the study was HIPAA compliant.

Study recruitment from the community commenced over a six-month period in 2021. Inclusion criteria were: 1) age of 21 years or older and 2) symptomatic knees associated with pain and/or a self-diagnosis of osteoarthritis. Exclusion criteria consisted of 1) orthopedic implants in the knee region, 2) other general contraindications for MRI, or 3) excessive motion in the scans.

The cohort included thirty-two participants, with the cohort size chosen based on the feasibility of radiologists completing the study with five protocols per participant.

### 2.2. Study Design and MRI Protocols

All subjects were scanned on a whole-body 3T MRI system (Signa Premier, GE Healthcare) using a medium-sized, flexible 16-channel receive-only phased-array-coil (Neocoil, Milwaukee, USA). The integrated body coil of the MRI system was used for excitation. All participants underwent the five protocols listed below in succession in one ∼40-minute scan session. All scan parameters are shown in Table 1.

**Table 1.** Overview of MRI Sequence Parameters for the Reference and Four Accelerated Knee MRI Protocols.

| Scan Protocol | Sequence | TA<br>(min<br>:sec) | TR<br>(ms) | TE<br>(ms) | FA<br>(°) | ETL | Slices | Acc<br>Phase | NEX | Matrix<br>size | FOV<br>(cm) | Voxel Size<br>(mm<sup>3</sup>) | (No)<br>Phase<br>Wrap | BW<br>(kHz) |
| --- | --- | --- | --- | --- | --- | --- | --- | --- | --- | --- | --- | --- | --- | --- |
| **Reference**<br>**Conventional**<br>**2D FSE**<br>**Protocol**<br>**[12-15**<br>**minutes]** | Axial T2<br>FS | 1:58 | 5588 | 61 | 111 | 14 | 31 | 2 | 1 | 320x256 | 16 | 0.5x0.6x3 | 2 | ±31.25 |
|  | Coronal<br>PD FS | 2:22 | 3555 | 30 | 111 | 6 | 37 | 2 | 1 | 320x288 | 16 | 0.5x0.6x3 | 1.5 | ±31.25 |
|  | Sagittal T2<br>FS | 2:09 | 4455 | 50 | 111 | 10 | 33 | 2 | 1 | 320x256 | 16 | 0.5x0.6x3 | 2 | ±31.25 |
|  | Sagittal<br>PD | 1:50 | 2500 | 22 | 111 | 6 | 33 | 2 | 1 | 384x312 | 16 | 0.4x0.5x3 | 1.5 | ±62.50 |
|  | Coronal<br>PD | 1:25 | 2507 | 27 | 111 | 6 | 37 | 2 | 1 | 320x288 | 16 | 0.5x0.6x3 | 1.2 | ±41.67 |
|  | Coronal<br>T1 | 1:38 | 789 | 9 | 111 | 3 | 37 | 2 | 1 | 416x224 | 16 | 0.4x0.7x3 | 1.5 | ±50 |
| **DL 2D FSE**<br>**Protocol**<br>**[6-7 minutes]** | Axial T2<br>FS | 0:51 | 4599 | 65 | 111 | 15 | 31 | 2 | 1 | 320x256 | 16 | 0.5x0.6x3 | 1 | ±62.50 |
|  | Coronal<br>PD FS | 1:15 | 3381 | 35 | 111 | 8 | 37 | 2 | 1 | 320x288 | 16 | 0.5x0.6x3 | 1 | ±62.50 |
|  | Sagittal T2<br>FS | 1:11 | 4150 | 50 | 111 | 13 | 33 | 2 | 1 | 320x256 | 16 | 0.5x0.6x3 | 1.5 | ±62.50 |
|  | Sagittal<br>PD | 1:15 | 2500 | 22 | 111 | 6 | 33 | 2 | 1 | 384x312 | 16 | 0.4x0.5x3 | 1 | ±83.33 |
|  | Coronal<br>PD | 0:45 | 2491 | 27 | 111 | 8 | 37 | 2.5 | 1 | 320x288 | 16 | 0.5x0.6x3 | 1 | ±83.33 |
|  | Coronal<br>T1 | 1:09 | 755 | 9 | 111 | 4 | 44 | 2 | 1 | 416x224 | 16 | 0.4x0.7x2.5 | 1.25 | ±50 |
| **3D qDESS**<br>**Protocol**<br>**[5-6 minutes]** | qDESS | 4:32 | 17 | 6,<br>29 | 20 | - | 150 | - | 1 | 384x350 | 16 | 0.4x0.4x1 | 1.5 | ±41.67 |
|  | Coronal<br>PD FS FSE | 1:15 | 3381 | 35 | 111 | 8 | 37 | 2 | 1 | 320x288 | 16 | 0.5x0.6x3 | 1 | ±62.50 |
|  | Cartilage<br>T2 Maps<br>(from<br>qDESS)<sup>8</sup> |  |  |  |  |  |  |  |  |  |  |  |  |  |
|  | Synovitis<br>Hybrid<br>Images<br>(from<br>qDESS)<sup>10</sup> |  |  |  |  |  |  |  |  |  |  |  |  |  |
| **3D Cube**<br>**Protocol**<br>**[5-6 minutes]** | Sagittal<br>PD CUBE | 5:04 | 1000 | 35 | 160 | 60 | 254 | 2 | 1 | 352x352 | 16 | 0.5x0.5x0.6 | 1 | ±62.50 |
|  | Coronal<br>PD FS FSE | 1:15 | 3381 | 35 | 111 | 8 | 37 | 2 | 1 | 320x288 | 16 | 0.5x0.6x3 | 1 | ±62.50 |
| **2D Thin-Slice**<br>**Protocol**<br>**[5 minutes]** | Sagittal<br>PD FS FSE | 3:54 | 6144 | 35 | 111 | 9 | 115 | 2.5 | 1 | 384x266 | 16 | 0.4x0.6x0.8 | 1.25 | ±62.50 |
|  | Coronal<br>T1 FSE | 1:09 | 755 | 9 | 111 | 4 | 44 | 2 | 1 | 416x224 | 16 | 0.4x0.7x2.5 | 1.25 | ±50 |

FSE Fast Spin Echo; FS fat suppressed; PD proton density; TA acquisition time; TR repetition time; TE echo time; FA flip angle; ETL echo train length; Acc acceleration factor; NEX number of excitations; FOV field of view; BW bandwidth.

1: **Reference conventional 2D FSE protocol:** This protocol included six accelerated sequences^4^, including axial T2-weighted fat-saturated (FS), coronal proton density (PD)-weighted FS, sagittal T2-weighted FS, sagittal PD, coronal PD, and coronal T1-weighted FSE images. This protocol is representative of a conventional clinical knee protocol that is commonly run for routine knee MRI at our center.

2: **2D DL FSE protocol:** This protocol included six accelerated 2D FSE sequences reconstructed using the GE AIR DL denoising reconstruction^6^. The same sequences as the conventional 2D FSE protocol were used with similar echo times, with modified parallel imaging acceleration, bandwidth, echo train lengths (ETL), and no-phase-wrap factors, to reduce scan times by roughly twofold.

3: **3D Cube protocol:** This protocol included a 3D PD-FSE sequence with variable flip-angle refocusing (Cube)^7^ and a 2D coronal PD-FS-FSE sequence with DL reconstruction for visualization of collateral ligaments and bone marrow lesions (BMLs), which are more apparent due to the fat-saturation and fluid sensitivity of this sequence. The SNR-efficiency of Cube allows for near-isotropic acquisitions and multi-planar reformatting in arbitrary planes.

4: **3D quantitative Double-Echo Steady-State (qDESS) protocol:** This protocol included a qDESS scan which offers multiple contrasts, and a 2D coronal PD-FS-FSE sequence with DL reconstruction for evaluation of BMLs and collateral ligaments. qDESS produces two echoes (S+ and S-), where S+ has high SNR with T1/T2 weighting while S- has fluid sensitivity with higher T2 weighting. The SNR-efficiency of qDESS allows for near-isotropic acquisitions and multi-planar reformatting in arbitrary planes. Additionally, analytical signal equations fit to the two echoes were used to create synovitis hybrid images (joint-fluid-suppressed images to provide contrast to synovial hypertrophy)^8^ and cartilage T2 relaxation time maps^9,10^.

5: **Thin-slice 2D FSE protocol:** This protocol included a 2D sagittal PD-FS-FSE sequence^11^ with a four-fold improvement in slice resolution as compared to the conventional scans and a 2D coronal T1-FSE sequence. This protocol has similar 2D FSE acquisitions at a lower slice thickness to allow for multiplanar reformats.

### 2.3. Image Analysis

The retrospectively constructed knee MRI protocols were independently evaluated by five radiologists, with five (R.V.H.), six (M.P.), ten (J.M.), seventeen (M.Y.), and thirty (K.S.) years of experience in interpreting clinical knee MRI examinations, including residency, at the start of the study. Each radiologist was blinded to patients and scan protocol and received five compilations of thirty-two randomly distributed datasets with a two-week washout period in between. The readers were provided with detailed instructions for image assessment, including example images and definitions of gradings. The ground truth radiologic finding was determined by taking the consensus (at least three out of five readers) of the reader scores for the reference conventional protocol. One radiologist (M.Y.) rescored the thirty-two reference clinical protocol scans two weeks after finishing the five compilations for repeatability analysis.

### 2.4. Outcome Measures

#### 2.4.1. Diagnostic Performance

Based on prior studies^3,12,13^, structures in the knee joints were assessed for the presence or absence of pathological findings. The findings were then binarized into non-pathologic (binarized scoring = 0) or pathologic (binarized scoring = 1), as described in Table 2.

**Table 2.** Overview of the Scoring Guide for Knee Tissue Derangements.

| **Tissue** | **Subregions** | **Original Scoring for Derangements** | **Binarized Scoring for Derangements** |
| --- | --- | --- | --- |
| Bone | Femur: medial condyle, lateral condyle, trochlea<br>Tibia: medial and lateral tibial plateau<br>Patella | 1: Bone marrow edema<br>Subchondral cyst<br>Subchondral sclerosis<br>2: Fracture | 1: Bone marrow edema<br>Subchondral cyst<br>Subchondral sclerosis<br>1 (Separate Analysis): Fracture |
| Ligaments | Collateral: MCL, LCL, PLC<br>Cruciate: ACL, PCL | 1: Sprain<br>2: Myxoid degeneration<br>Partial tear<br>3: Complete tear | 0: Sprain<br>1: Myxoid degeneration<br>Partial tear<br>Complete tear |
| Synovium |  | Grades 0-3: Joint effusion<br>Grades 0-3: Synovitis | 0: Grades 0, 1<br>1: Grades 2, 3 (for both) |
| Cartilage | Femur: medial condyle, lateral condyle, trochlea<br>Tibia: medial and lateral tibial plateau<br>Patella | 1: Increased T2 signal of normal cartilage<br>2A: Superficial partial thickness defect <50%<br>2B: Deep partial thickness defect >50%<br>3: Full thickness chondral defect | 0: Increased T2 signal of normal cartilage<br>1: Superficial partial thickness defect <50%<br>Deep partial thickness defect >50%<br>Full thickness chondral defect |
| Meniscus | Medial and lateral | 1: Myxoid degeneration<br>2: Horizontal tear<br>3: Radial tear<br>4: Vertical longitudinal tear<br>5: Complex<br>6: Complex flap | 0: Myxoid degeneration<br>1: Horizontal tear<br>Radial tear<br>Vertical longitudinal tear<br>Complex<br>Complex flap |

ACL anterior cruciate ligament; PCL posterior cruciate ligament; MCL medial collateral ligament; LCL lateral collateral ligament; PLC posterolateral corner.

#### 2.4.2. Repeatability Analysis

One radiologist rescored the thirty-two reference clinical protocol scans for repeatability analysis.

#### 2.4.3. Image Diagnostic Quality

A visual assessment of diagnostic image quality was performed between protocol groups. The degree of artifacts and general image quality were assessed using a five-point Likert-type scale. A score of 1 (“completely non-diagnostic”) indicated complete obscuration of anatomic details and imaging findings, a score of 2 (“severe corruptions”) indicated obscuration of most anatomic details and imaging, a score of 3 (“diagnostically acceptable”) indicated obscuration of fine anatomic details, a score of 4 (“good quality”) indicated minimal obscuration of fine anatomic details, and a score of 5 (“very good quality”) indicated full visibility of anatomic details.

#### 2.4.4. Utility of 3D Reformats

Beyond diagnostic accuracy and confidence, we evaluated the utility of oblique image reformats in 3D qDESS and Cube, and 2D Thin-Slice protocols. Readers were asked to rate whether 3D reformats were useful on a scale from 0 to 3, where 1- “helpful in validating diagnosis”, 2- “helpful in making a diagnosis”, and 3- “would not have made a diagnosis without them”.

### 2.5. Statistical Analysis

A biostatistician (T.L.) performed the statistical analyses using Stata Statistical Software (version 18, StataCorp LLC).

#### 2.5.1. Diagnostic Performance

The five readers graded structural abnormalities of the knee as either present or absent, spanning menisci, ligaments, bone marrow, cartilage, synovitis and effusion. Inter-reader and inter-method concordance agreements (four approaches vs reference protocol) were quantified using Gwet’s AC. Diagnostic performances were calculated as sensitivity and specificity.

#### 2.5.2. Repeatability Analysis

Repeatability analysis was performed using Gwet’s AC for pathology scoring.

#### 2.5.3. Image Diagnostic Quality

Image quality Likert scale grades were reported as the mean and standard deviation values, along with the percentage of grades greater than 3 (“diagnostically acceptable”). Differences between image quality evaluations for various tissues and readers were assessed with the Friedman test. *p* < .05 was considered a statistically significant difference.

#### 2.5.4. Utility of 3D Reformats

The scores ranged from 0-3 and were presented as a percentage of the total number of scores for each protocol-Cube, qDESS and Thin-Slice.

## 3. Results

### 3.1. Participant Characteristics

32 participants were enrolled in the study: 14 male and 18 female (average age 47 ± 16 years, other information in Table 3). Some image sets had motion artifacts that resulted in them being non-diagnostic and they were therefore removed from the analysis. Of the 32 subjects, 1 subject was excluded due to motion artifacts in 3 out of 5 protocols; out of the remaining 155 scans (5 scans per participant), 7 scan protocols were excluded due to severe motion. Figure 1 displays the flow diagram for the study.

**Figure 1.**
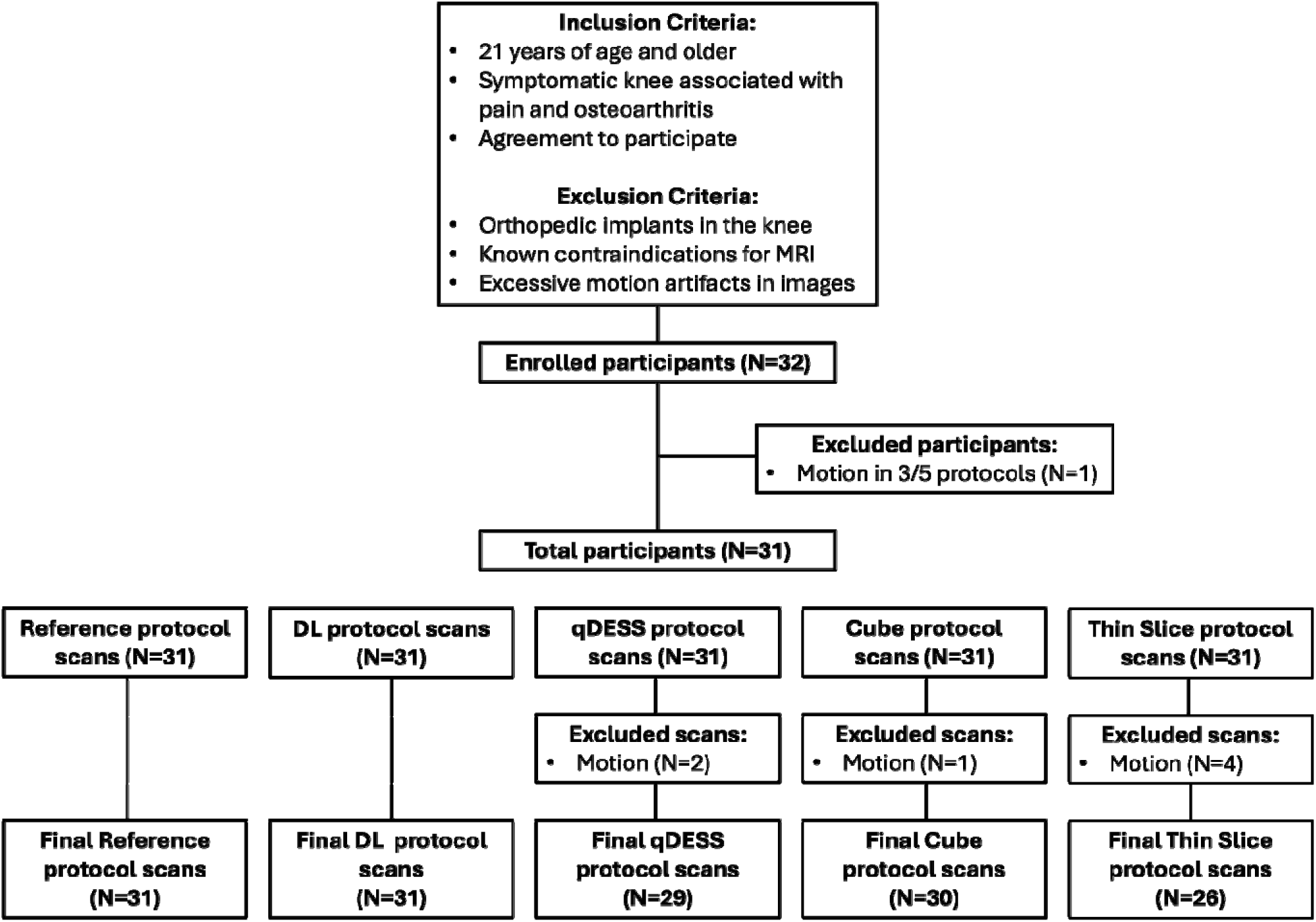
Flow diagram shows participants and protocols in the study.

**Table 3.** Patient demographics.

| Characteristics | Results (N=31) |
| --- | --- |
| Age (years)* | 23-78 (48 ± 16) |
| Sex |  |
| Female | 13 (42%) |
| Male | 18 (58%) |
| BMI (kg/m<sup>2</sup>)* | 18-36 (26 ± 4) |
| Laterality |  |
| Right Knee | 17 (56%) |
| Left Knee | 14 (45%) |

\* Data represents a range, with means ± SDs in parentheses.

### 3.2. Diagnostic Performance

Figures 2-5 show imaging results for a male participant aged 31-35 years with knee pain and the following findings: lateral tibial bone marrow edema, ACL reconstruction, femoral and tibial cartilage damage, osteophytes in the femur, tibia, and patella, and abnormal lateral meniscus morphology. Figure 2 compares Conventional 2D FSE and Accelerated 2D FSE with Deep Learning (DL) Reconstruction, with DL providing superior image quality while equally capturing pathologies. Figure 3 shows Cube protocol images, offering similar pathology detection as 2D FSE but with the advantage of multiplanar reformats. Figure 4 presents qDESS protocol images, including multiplanar echo 1 and echo 2 views, synovitis hybrid images, and T2 cartilage relaxation time maps. Figure 5 highlights the Thin-Slice protocol images with and without DL reconstruction, with DL reconstruction enhancing image quality by denoising and reducing blocky patterns in multiplanar reformats.

**Figure 2.**
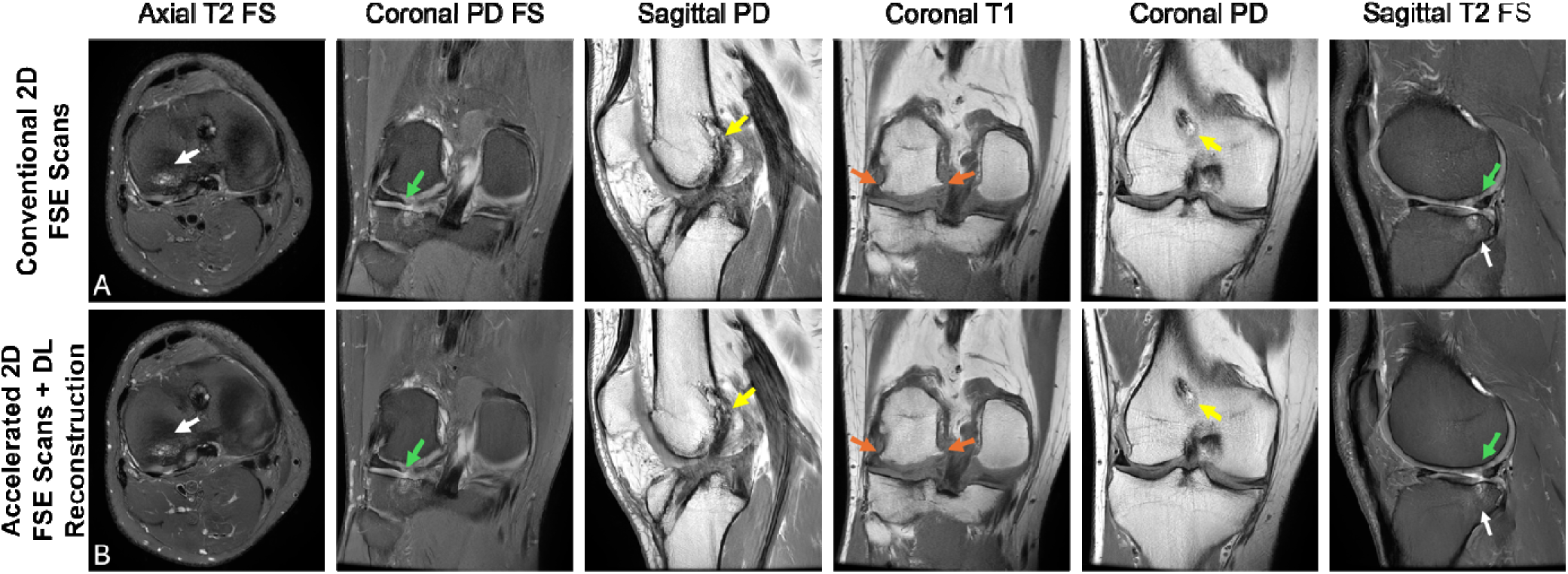
A) Conventional 2D FSE and B) Accelerated 2D FSE Protocol with Deep Learning (DL) Reconstruction. A male participant aged 31-35 years with knee pain presents with lateral tibial bone marrow edema (white arrows), ACL reconstruction (yellow arrows), lateral and medial femoral and tibial cartilage damage, osteophytes (orange arrows), and abnormal morphology of posterior horn of the lateral meniscus likely due to either previous partial meniscectomy or degenerative partial maceration (green arrows). The pathologic features can be seen equally well in both protocols. Each column represents the images acquired in multiple planes, with multiple contrasts. [FSE Fast Spin Echo; FS fat suppressed; PD proton density].

**Figure 3.**
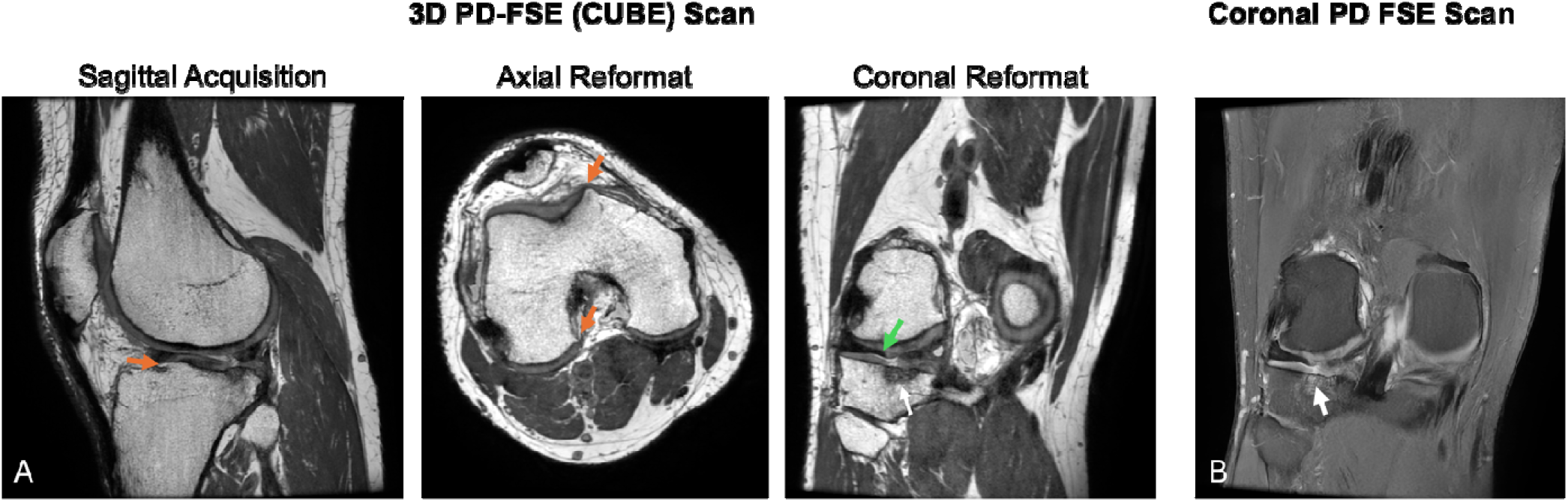
The CUBE protocol, including a A) 3D sagittal PD-FSE (CUBE) scan and a B) coronal PD FSE scan. The CUBE scan included coronal and axial reformats. Some pathological include lateral tibial bone marrow edema (white arrows), lateral and medial femoral and tibial cartilage damage, osteophytes (orange arrows), and abnormal morphology of posterior horn of the lateral meniscus (green arrow). [FSE Fast Spin Echo; FS fat suppressed; PD proton density].

**Figure 4.**
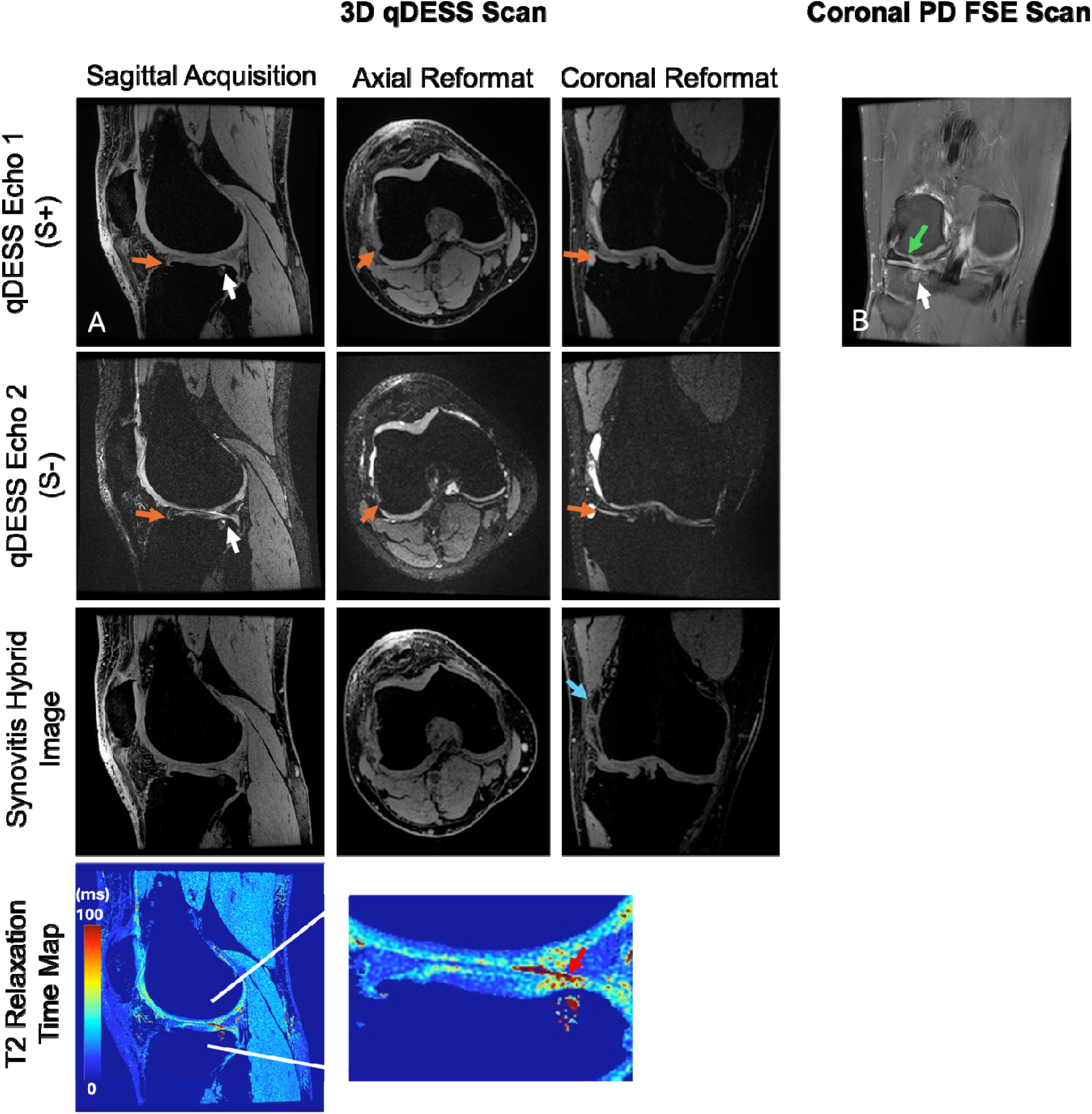
The quantitative double-echo steady state (qDESS) protocol, including A) 3D qDESS scan and B) coronal PD FS FSE scan. The qDESS images (echo 1 and echo 2) were used to create synovitis hybrid images and T2 relaxation time maps, and included coronal and axial reformats. The zoomed in images show cartilage damage in the lateral tibial region (red arrow), with changes in T2 relaxation times observed in the corresponding T2 map, located near corresponding bone marrow edema observed in the PD FSE scan (white arrows). The qDESS scans were also helpful for identifying osteophytes (orange arrows) and fluid (blue arrow).

**Figure 5.**
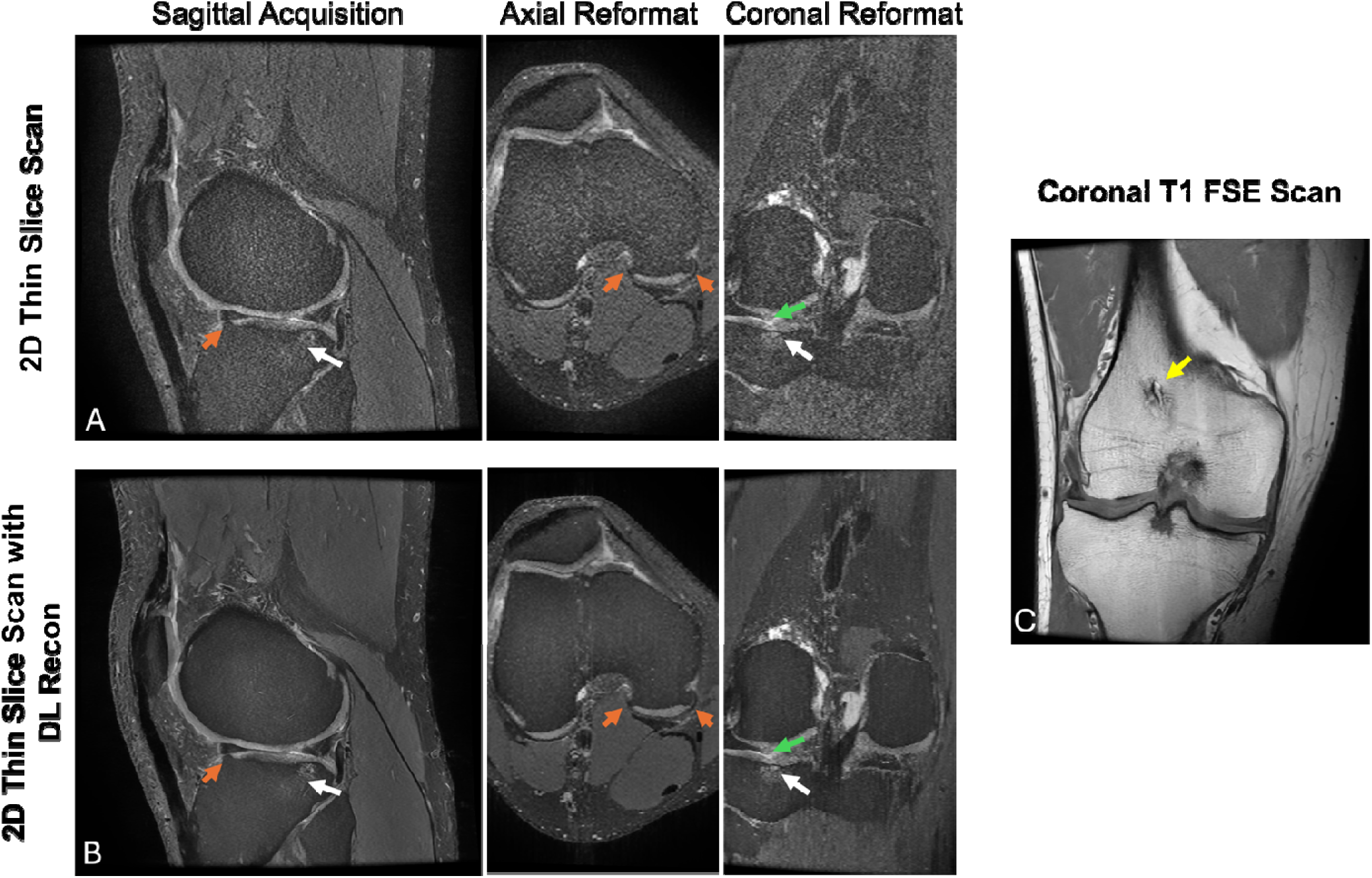
Thin Slice protocol, with and without DL reconstruction. This protocol included a A) 2D sagittal PD-FS-FSE (Thin-Slice) scan, a B) 2D sagittal PD-FS-FSE (Thin-Slice) scan with DL Reconstruction and a C) coronal T1 FSE scan. The Thin Slice scan included coronal and axial reformats. Observed pathological findings include lateral tibial bone marrow edema (white arrows), lateral and medial femoral and tibial cartilage damage, osteophytes (orange arrows), and abnormal morphology of posterior horn of the lateral meniscus (green arrows). Both protocols can detect pathological findings, but the higher image quality with DL reconstruction enhances the diagnostic utility, especially for the axial and coronal reformats. [FSE Fast Spin Echo].

Table 4 summarizes the inter-reader and within-reader inter-method agreement, sensitivity, and specificity analysis of the accelerated knee MRI protocols with respect to the reference protocol. ACL pathology results were not reported as only 2 instances of pathology were observed, making statistical analysis impossible.

**Table 4.** Diagnostic performance of the four accelerated protocols in comparison with the reference clinical protocol.

| Measure | Protocol | Bone Marrow | Cartilage | Meniscus | Synovitis | Effusion |
| --- | --- | --- | --- | --- | --- | --- |
| **Inter-reader Agreement** | **Reference** | 0.79<br>(0.71, 0.90) | 0.45<br>(0.41, 0.52) | 0.74<br>(0.73, 0.75) | 0.83 | 0.85 |
|  | **2D DL** | 0.75<br>(0.62, 0.86) | 0.41<br>(0.35, 0.46) | 0.83<br>(0.80, 0.85) | 0.80 | 0.89 |
|  | **3D Cube** | 0.76<br>(0.69, 0.89) | 0.41<br>(0.25, 0.53) | 0.83<br>(0.82, 0.84) | 0.78 | 0.87 |
|  | **3D qDESS** | 0.80<br>(0.74, 0.88) | 0.48<br>(0.40, 0.54) | 0.86<br>(0.83, 0.88) | 0.65 | 0.80 |
|  | **2D Thin-Slice** | 0.77<br>(0.71, 0.84) | 0.44<br>(0.32, 0.56) | 0.69<br>(0.63, 0.76) | 0.87 | 0.87 |
| **Inter-method Agreement** | **2D DL** | 0.87<br>(0.56, 1.00) | 0.74<br>(0.34, 1.00) | 0.81<br>(0.62, 1.00) | 0.84<br>(0.76, 0.92) | 0.93<br>(0.88, 1.00) |
|  | **3D Cube** | 0.87<br>(0.63, 1.00) | 0.73<br>(0.35, 0.96) | 0.81<br>(0.61, 1.00) | 0.81<br>(0.63, 1.00) | 0.91<br>(0.87, 1.00) |
|  | **3D qDESS** | 0.85<br>(0.50, 1.00) | 0.77<br>(0.41, 1.00) | 0.84<br>(0.62, 1.00) | 0.74<br>(0.61, 0.89) | 0.85<br>(0.67, 1.00) |
|  | **2D Thin-Slice** | 0.83<br>(0.57, 1.00) | 0.64<br>(0.34, 0.83) | 0.71<br>(0.47, 0.86) | 0.84<br>(0.74, 1.00) | 0.89<br>(0.79, 0.96) |
| **Sensitivity (%)** | **2D DL** | 78<br>(50, 100) | 83<br>(40, 100) | 95<br>(77, 100) | 66<br>(29, 86) | 93<br>(67, 100) |
|  | **3D Cube** | 76<br>(25, 100) | 78<br>(21, 100) | 91<br>(75, 100) | 53<br>(33, 100) | 64<br>(20, 100) |
|  | **3D qDESS** | 60<br>(22, 100) | 73<br>(13, 100) | 89<br>(73, 100) | 67<br>(33, 83) | 80<br>(40, 100) |
|  | **2D Thin-Slice** | 72<br>(0, 100) | 74<br>(42, 100) | 79<br>(70, 90) | 44<br>(40, 100) | 60<br>(20, 80) |
| **Specificity (%)** | **2D DL** | 91<br>(69, 100) | 78<br>(25, 100) | 93<br>(73, 100) | 93<br>(88, 100) | 92<br>(88, 100) |
|  | **3D Cube** | 86<br>(0, 100) | 81<br>(39, 100) | 94<br>(88, 100) | 93<br>(88, 100) | 96.8<br>(92, 100) |
|  | **3D qDESS** | 93<br>(77, 100) | 86<br>(38, 100) | 98<br>(88, 100) | 88<br>(78, 96) | 92<br>(83, 100) |
|  | **2D Thin-Slice** | 89<br>(71, 100) | 78<br>(27, 100) | 88<br>(73, 100) | 93<br>(91, 100) | 96<br>(95, 100) |
| **Repeatability** | **Reference** | 0.89<br>(0.75, 0.96) | 0.81<br>(0.68, 0.89) | 0.91<br>(0.88, 0.95) | 0.84 | 0.95 |

Data in parentheses shows the range of values for all subregions and readers. For synovitis and effusion, only whole pathologies were analyzed. Inter-reader agreement, inter-method agreement, and repeatability data are Gwet's AC values.

For bone marrow pathology, the 3D qDESS protocol demonstrated the highest inter-reader agreement (0.80), followed by the 3D CUBE protocol (0.76), while the inter-method agreement was slightly higher for 2D DL and 3D CUBE (both 0.87) protocols. The 2D DL protocol had the highest sensitivity (78%), but specificity was highest for 3D qDESS (93%).

For cartilage pathology, we observed that the 3D qDESS protocol showed the highest inter-reader agreement (0.48), inter-method agreement (0.77), and specificity (86%). Sensitivity was highest for 2D DL (83%).

For meniscus pathology, the 3D qDESS protocol achieved the highest inter-reader agreement (0.86), inter-method agreement (0.84), sensitivity (95%), and specificity (98%). Excellent reader agreement is observed for meniscus evaluations, particularly with 3D qDESS, with similar performances seen for the remaining 3 protocols.

For synovitis, the 2D Thin Slice protocol had the highest inter-reader agreement (0.87) and inter-method agreement (0.84). Sensitivity was highest for 2D DL (66%), while specificity was comparable across methods, with all methods exceeding 88%.

Lastly, for effusion, both 2D DL and 3D CUBE protocols showed strong performance, with high inter-reader agreement (0.89 and 0.87, respectively). Specificity was highest for 3D CUBE (96.8%), while 2D DL had the highest sensitivity (93%).

Overall, the 3D qDESS emerged as the most consistent method for imaging the meniscus and cartilage, with the 2D DL and 3D CUBE protocols showing strong performances for bone marrow pathology and effusion. Comparatively, these three protocols performed equivalently across the different pathologies. While the 2D Thin-Slice protocol showed promise for synovitis imaging, it did not perform as well for other pathologies.

### 3.3. Repeatability

Overall, the repeatability between Reader M.Y.’s scores for the reference protocol at two time points (original reader study and repeated rescoring) was excellent across pathologies, with cartilage showing the lowest Gwet’s AC values (0.68-0.89) and effusion with the highest (0.95). The other pathologies showed Gwet’s AC values ranging from 0.84-0.89 (Table 4).

### 3.4. Image Diagnostic Quality

The overall image diagnostic quality for all five protocols ranged from score 1 (“diagnostically acceptable”) to score 5 (“very good quality”) for all pathological features (Figure 6). The results are shown for four different anatomical regions: bone marrow, cartilage, meniscus, and ligaments. A score of 3 was considered diagnostically acceptable, and for all five approaches for the bone marrow, meniscus, and ligaments, the diagnostic quality scores are above this level across all readers for more than 95% of all measurements. For cartilage, the diagnostic quality scores are above 3 for 80.5% to 99.1% of all measurements.

**Figure 6.**
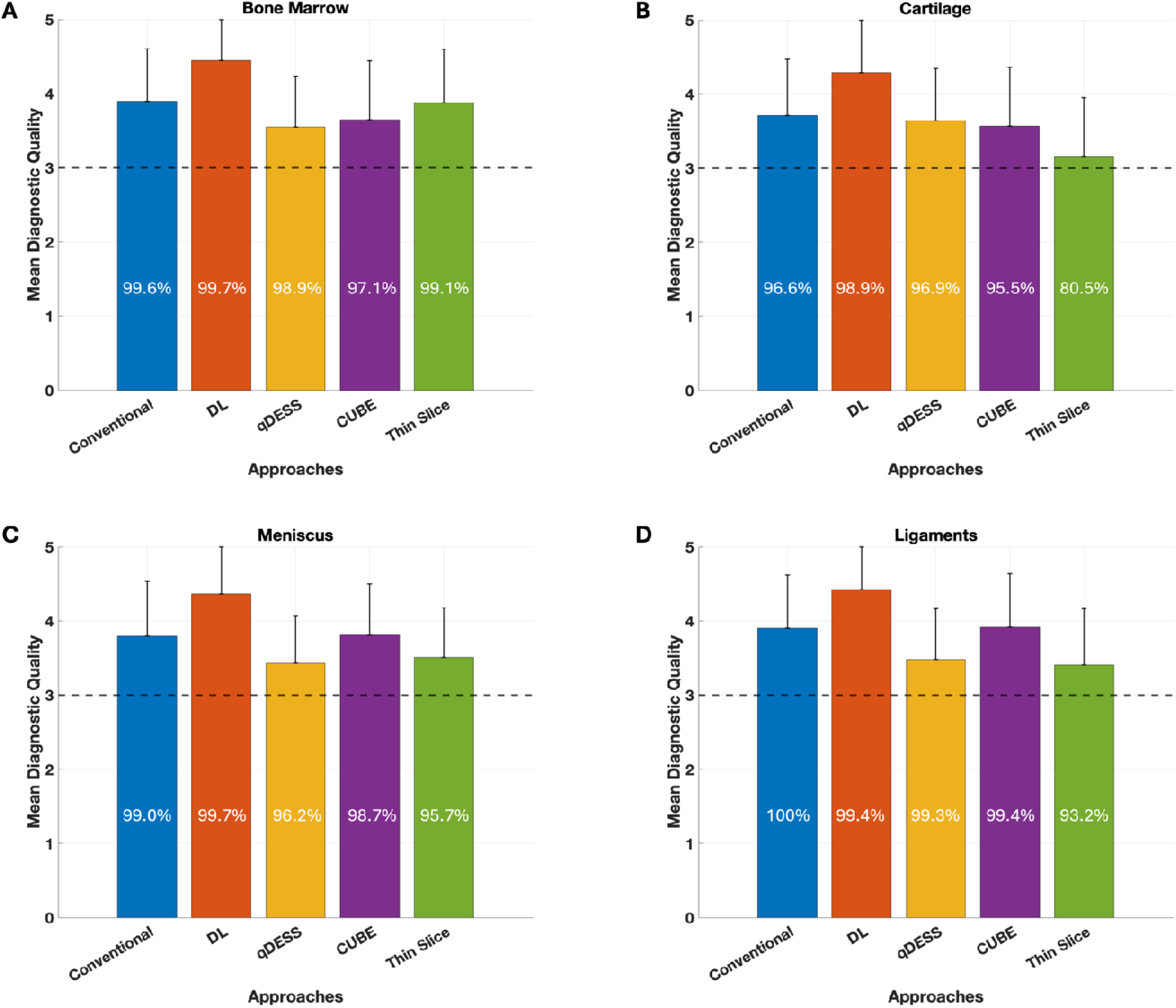
The bar graphs show results for the mean image diagnostic quality scores for the five protocols: Conventional, DL, qDESS, Cube, and Thin Slice, for four tissue groups: A) Bone Marrow, B) Cartilage, C) Meniscus, and D) Ligaments. Scores are averaged over all data points, across all five readers. The scoring ranged from 1 (“Completely Non-Diagnostic”) to 5 (“Very Good Quality”). All protocols for all tissue groups showed image quality more than the “Diagnostically Acceptable” level, shown using a horizontal line drawn at score 3 out of 5, and the percentages represent the number of measurements scored greater than equal to 3 out of all measurements.

### 3.5. Utility of 3D Reformats

For the qDESS protocol, 3D reformats were helpful in 74% of cases (56% in validating diagnosis, 14% in making diagnosis, and 4% vital in diagnosis – i.e. diagnosis would not have been made without). Similarly, for the Cube protocol, readers found 3D reformats helpful in 74% of the cases (41% in validating diagnosis, 25% in making diagnosis, and 8% vital in diagnosis). Lastly, for the thin-slice protocol, the 3D reformats were helpful in 69% of the cases (52% in validating diagnosis, 7% in making diagnosis, and 10% vital in diagnosis).

## 4. Discussion

This study evaluated four accelerated knee MRI protocols (∼6 minutes each) against a standard clinical protocol (15-30 minutes) across various knee pathologies. Overall, three protocols— accelerated conventional protocol with deep learning (DL) reconstruction, Cube, and qDESS— demonstrated similar inter-reader and inter-method agreement to the conventional protocol, with slightly lower agreement observed for the Thin-Slice protocol. The diagnostic quality of DL, Cube, and qDESS was deemed diagnostically acceptable or superior for all pathologies, suggesting their potential to replace conventional protocols and significantly reduce scan times.

Reader interpretations about what is considered ‘pathological’ varied, particularly for cartilage pathology, matching prior studies showing variability in cartilage lesion scoring^8,14^. Two reviewers were more conservative in identifying pathology, which slightly lowered sensitivity when compared to the consensus reference standard (3 out of 5 readers’ scores). Despite this, within-reader scoring of pathology remained consistent across protocols, especially for the DL, Cube, and qDESS protocols, relative to the reference protocol. These three protocols showed inter-reader and within-reader inter-method agreements comparable to or better than the conventional protocol, reinforcing their diagnostic equivalence to the current clinical convention.

The **2D accelerated conventional protocol with DL reconstruction** provided high-quality images despite a two-factor acceleration and reduction in scan time, with noise reduction and motion artifact correction contributing to sharp, high-contrast images. Diagnostic quality was rated similar to or better than the conventional protocol, aligning with prior studies^2,3,15^. A big advantage of this protocol is its use of contrasts and sequences similar to conventional protocols, ensuring its adaptability and ease of clinical implementation.

The **3D qDESS protocol** offered high signal-to-noise ratio (SNR) efficiency and simultaneous morphological and quantitative image information. Our study, which utilized blinded assessments with washout periods, supports previous findings of near-perfect agreement between this protocol and the conventional approach observed in direct side-by-side comparisons^8–10^. This protocol offers oblique reformats due to its high through-plane resolution and the possibility of DL reconstruction. Furthermore, new information such as synovitis hybrid images for detecting synovial hypertrophy^8^, and cartilage T2 maps for identifying early microstructural changes^10^, can impact clinical evaluation of early osteoarthritis and effects of future disease-modifying osteoarthritis drugs.

The **3D Cube protocol** provided improved resolution, high SNR efficiency, and oblique reformats with a contrast similar to the conventional protocol^7^. At the time of study initiation (2021), a non-fat saturated 3D Cube approach was chosen due to some blurring effects in fat-saturated 3D Cube. However, advances in acquisition speed allow for fat-saturated T2-weighted 3D Cube with improved image quality and reduced through-slice blurring. Future iterations, combining advanced Cube sequences with DL reconstruction, may further enhance diagnostic performance, despite the protocol already demonstrating equivalence to the conventional approach.

The **2D Thin-Slice protocol** delivered promising results but lagged behind the other three protocols. This protocol offered an opportunity to push the limits of DL reconstruction, allowing familiar 2D acquisitions at a high slice thickness to allow for reformats^11^. However, increased slice resolution reduced SNR, and DL reconstruction effects were primarily in-plane, leading to blocky patterns in reformats. Future iterations, such as a 3D thin-slice approach with DL reconstruction, may improve its performance and SNR efficiency.

Modern MRI technologies, including novel 3D sequences, reconstruction methods, and hardware advances, enable accelerated imaging without compromising diagnostic quality. Simultaneous multi-slice (SMS), parallel imaging, and compressed sensing techniques significantly accelerate MRI acquisition, achieving 2-fold faster 2D FSE imaging^16–18^ and 3-to 5-fold faster 3D sequences like SPACE TSE and Cube FSE^19–21^, while maintaining diagnostic quality. Deep learning is a powerful tool for reconstructing high-quality, accelerated MR images from undersampled data^2,3^, enhancing resolution, reducing noise, correcting artifacts^23^, and creating synthetic contrasts^24^. DL applications continue to expand, improving 3D sequences and the entire imaging pipeline from acquisition to diagnosis. Hardware innovations like novel flexible coils (AIR coils, High-impedance coils (HIC)) and bilateral coil arrays^25^ have further improved SNR and reduced acquisition times, without loss of diagnostic quality. Furthermore, as reducing MRI acquisition time doesn’t always shorten scan slots, strategies like employing additional staff, dockable tables, and optimized facility design, can help decrease outpatient wait times^26–27^, highlighting the potential of combining imaging advances with optimized clinical workflows to improve patient care.

Our study had some notable limitations. The study’s small sample size limits generalizability, and the absence of a gold standard reference constrains objective diagnostic accuracy assessments. The use of a state-of-the-art scanner and optimized coils may not directly translate to all systems, particularly for protocols like Cube, which are sensitive to gradient strength. Additionally, binarized scoring of pathologies simplified radiological reporting but sacrificed detail, warranting future studies exploring semi-quantitative scoring methods like MOAKS and BLOKS for osteoarthritis. Furthermore, some datasets, primarily in the qDESS, Cube, and Thin-Slice protocols, were excluded due to motion artifacts. This was likely due to the sequences being performed later in the protocol, leading to subject fatigue and increased movement.

In conclusion, three accelerated knee imaging protocols—DL, Cube, or qDESS—have the potential to replace the conventional knee MRI protocol and dramatically reduce scan times, while maintaining diagnostic performance and image quality. These protocols can reduce imaging times to less than six minutes, enhancing patient throughput and comfort while offering consistent diagnostic performance for diverse knee pathologies. Tailored protocol selection could further optimize outcomes, paving the way for efficient and precise musculoskeletal imaging.

## Data Availability

All data produced in the present study are available upon reasonable request to the authors

## Abbreviations

2D: Two-dimensional
FSE: Fast Spin Echo
PD: Proton Density-weighted
3D: Three-dimensional
SNR: Signal-to-Noise Ratio
qDESS: quantitative Double-Echo Steady-State
DL: Deep Learning
FS: Fat Saturated

## References

1. American College of Radiology. ACR Practice Parameters and Technical Standards. Available at https://www.acr.org/Clinical-Resources/Practice-Parameters-and-Technical-Standards. Accessed January 5, 2021.

2. Recht MP, Zbontar J, Sodickson DK, et al. Using Deep Learning to Accelerate Knee MRI at 3 T: Results of an Interchangeability Study. AJR Am J Roentgenol. 2020;215(6):1421–1429. doi:10.2214/AJR.20.23313

3. Johnson, P. M., Lin, D. J., Zbontar, et al. (2023). Deep Learning Reconstruction Enables Prospectively Accelerated Clinical Knee MRI. Radiology, 307(2), Article e220425. 10.1148/radiol.220425

4. Chaudhari AS, Kogan F, Pedoia V, Majumdar S, Gold GE, Hargreaves BA. Rapid knee MRI acquisition and analysis techniques for imaging osteoarthritis. J Magn Reson Imaging 2020; 52:1321–1339

5. Iuga, A. I., Abdullayev, N., Weiss, K., Haneder, S., Brüggemann-Bratke, L., Maintz, D., Rau, R., & Bratke, G. (2020). Accelerated MRI of the knee. Quality and efficiency of compressed sensing. European journal of radiology, 132, 109273. 10.1016/j.ejrad.2020.109273

6. Peters RD, Harris H, Lawson S (2020) The clinical benefits of AIR™ Recon DL for MR image reconstruction. https://www.gehealthcare.com/ensg/-/jssmedia/c943df5927a049bb9ac95a9f0349ad8c.pdf. Accessed January 5, 2021.

7. Li CQ, Chen W, Rosenberg JK, et al. Optimizing isotropic three-dimensional fast spin-echo methods for imaging the knee. J Magn Reson Imaging. 2014;39(6):1417–1425. doi:10.1002/jmri.24315.

8. Chaudhari AS, Stevens KJ, Sveinsson B, et al. Combined 5-minute double-echo in steady-state with separated echoes and 2-minute proton-density-weighted 2D FSE sequence for comprehensive whole-joint knee MRI assessment. J Magn Reson Imaging 2019;49(7):e183–e194.

9. Chaudhari AS, Grissom MJ, Fang Z, et al. Diagnostic Accuracy of Quantitative Multicontrast 5-Minute Knee MRI Using Prospective Artificial Intelligence Image Quality Enhancement. AJR Am J Roentgenol. 2021;216(6):1614–1625. doi:10.2214/AJR.20.24172.

10. Thoenen J, Stevens KJ, Turmezei TD, et al. Non-contrast MRI of synovitis in the knee using quantitative DESS. Eur Radiol. 2021;31(12):9369–9379. doi:10.1007/s00330-021-08025-2.

11. Tokuda O, Harada Y, Shiraishi G, et al. MRI of the anatomical structures of the knee: the proton density-weighted fast spin-echo sequence vs the proton density-weighted fast-recovery fast spin-echo sequence. Br J Radiol. 2012;85(1017):e686–e693. doi:10.1259/bjr/99570113

12. Chaudhari AS, Grissom MJ, Fang Z, et al. Diagnostic Accuracy of Quantitative Multi-Contrast 5-Minute Knee MRI Using Prospective Artificial Intelligence Image Quality Enhancement. AJR Am J Roentgenol 2020. 10.2214/AJR.20.24172. Published online August 5, 2020. Accessed March 4, 2024.

13. Hunter, D. J., Guermazi, A., Lo, G. H., Grainger, A. J., Conaghan, P. G., Boudreau, R. M., & Roemer, F. W. (2011). Evolution of semi-quantitative whole joint assessment of knee OA: MOAKS (MRI Osteoarthritis Knee Score). Osteoarthritis and cartilage, 19(8), 990–1002. 10.1016/j.joca.2011.05.004.

14. Wong, S., Steinbach, L., Zhao, J., Stehling, C., Ma, C. B., & Link, T. M. (2009). Comparative study of imaging at 3.0 T versus 1.5 T of the knee. Skeletal radiology, 38(8), 761–769. 10.1007/s00256-009-0683-0.

15. Del Grande F, Rashidi A, Luna R, Delcogliano M, Stern S, Dalili FJD. “Five-Minute Five-Sequence Knee MRI Using Combined Simultaneous Multislice and Parallel Imaging Acceleration: Comparison with 10-Minute Parallel Imaging Knee MRI.” Radiology. Radiological Society of North America vol. 299,3 (2021): 635–646. doi:10.1148/radiol.2021203655

16. Gao, F., Wen, Z., Dou, S., Kan, X., Wei, S., & Ge, Y. (2021). High-Resolution Simultaneous Multi-Slice Accelerated Turbo Spin-Echo Musculoskeletal Imaging: A Head-to-Head Comparison With Routine Turbo Spin-Echo Imaging. Frontiers in physiology, 12, 759888. 10.3389/fphys.2021.759888.

17. Kreitner KF, Romaneehsen B, Krummenauer F, Oberholzer K, Muller LP, Duber C. Fast magnetic resonance imaging of the knee using a parallel acquisition technique (mSENSE): A prospective performance evaluation. Eur Radiol 2006; 16(8): 1659–1666.

18. Niitsu M, Ikeda K. Routine MR examination of the knee using parallel imaging. Clin Radiol 2003; 58(10): 801–807.

19. Notohamiprodjo M, Horng A, Pietschmann MF, et al. MRI of the knee at 3T: First clinical results with an isotropic PDfs-weighted 3D-TSE-sequence. Invest Radiol 2009; 44(9): 585–597.

20. Kijowski R, Davis KW, Woods MA, et al. Knee joint: comprehensive assessment with 3D isotropic resolution fast spin-echo MR imaging—diagnostic performance compared with that of conventional MR imaging at 3.0 T. Radiology 2009;252(2):486–495.

21. Lee SH, Lee YH, Suh JS. Accelerating knee MR imaging: Compressed sensing in isotropic three-dimensional fast spin-echo sequence. Magn Reson Imaging 2018; 46: 90–97.

22. Chaudhari AS, Fang Z, Kogan F, et al. Super-resolution musculoskeletal MRI using deep learning. Magn Reson Med 2018; 80(5): 2139–2154.

23. Kustner T, Armanious K, Yang J, Yang B, Schick F, Gatidis S. Retrospective correction of motion-affected MR images using deep learning frameworks. Magn Reson Med 2019; 82(4): 1527–1540.

24. Nykanen O, Nevalainen M, Casula V, et al. Deep-learning-based contrast synthesis from MRF parameter maps in the knee joint. J Magn Reson Imaging 2023; 58(2): 559–568.

25. Kogan F, Levine E, Chaudhari AS, et al. Simultaneous bilateral-knee MR imaging. Magn Reson Med 2018; 80(2): 529–537.

26. GharehMohammadi, F., & Sebro, R. A. (2024). Efficient Health Care: Decreasing MRI Scan Time. Radiology. Artificial intelligence, 6(3), e240174. 10.1148/ryai.240174

27. Recht, M. P., Block, K. T., Chandarana, H., Friedland, J., Mullholland, T., Teahan, D., & Wiggins, R. (2019). Optimization of MRI Turnaround Times Through the Use of Dockable Tables and Innovative Architectural Design Strategies. AJR. American journal of roentgenology, 212(4), 855–858. 10.2214/AJR.18.20459

28. Knoll F, Zbontar J, Sriram A, et al. fastMRI: A Publicly Available Raw k-Space and DICOM Dataset of Knee Images for Accelerated MR Image Reconstruction Using Machine Learning. Radiol Artif Intell. 2020;2(1):e190007. Published 2020 Jan 29. doi:10.1148/ryai.2020190007

29. Muckley MJ, Riemenschneider B, Radmanesh A, et al. Results of the 2020 fastMRI Challenge for Machine Learning MR Image Reconstruction. IEEE Trans Med Imaging. 2021;40(9):2306–2317. doi:10.1109/TMI.2021.3075856

